# Application of chaos and systems theories to explore health workforce re-organisation following district splitting in Uganda

**DOI:** 10.1101/2024.08.29.24312785

**Authors:** Aloysius Mutebi, Moses Mukuru, Suzanne Kiwanuka, Fredrick Makumbi, Elizeus Rutebemberwa

**Author notes:** **Corresponding author’ email address:**. **Email addresses of co-authors** Moses Mukuru Suzanne Kiwanuka Fredrick Makumbi Elizeus Rutebemberwa.

## Abstract

**Background:** District splitting (DS) began in 1962 and intensified in 1997 during implementation of decentralization. This legislative process focuses on geographical demarcation. The health system, organized along local government structures, was re-organized as a result of DS. However, this study explored how the health workforce, as a component of the local government health system, re-organized following district splitting.

**Methods:** Conducted between June and December 2018, the qualitative study involved 22 key informant interviews in eight districts, representing four broader regions (North, Western, Eastern and Central) of Uganda. Participants included district political leaders, local government staff, district service commission members, district health team officials, and district senior management. Data was collected using an interview guide and four themes were identified for deductive thematic analysis.

**Results:** Findings reveal that district splitting led to an involuntary division of the health system between old and new districts, triggering multidirectional reorganization of the health workforce. Health workers were systematically allocated to districts based on the geographical location of their health facilities during the splitting process. Subsequently, inter-district migration occurred through health workforce secondment or appointment, while intra-health system migration involved vertical movement of health workers within districts during recruitment. This reorganization resulted in workforce shortages in health facilities vacated by migrating health workers.

**Conclusions:** The study concludes that district splitting caused unintended consequences as a result of chaotic occurrences that led to reorganization of the health workforce in both new and old districts, and caused gaps in service delivery capabilities. Adopting a systems and chaos perspective, the study emphasizes the importance of considering frontline service delivery systems as subsystems of the overall district system when evaluating the impact of district splitting. Such an approach provides a comprehensive framework for understanding the disruptive effects of district splitting on health system organization and service delivery.

## 1.0 Introduction

Decentralized administrative units are crucial for effective service delivery worldwide (Walker & Andrews, 2015). In Uganda, the current local government structures can be traced back to 1948, with district demarcation beginning in 1959. District splitting and subsequent, creation of new districts, is part of decentralization efforts to bring services closer to citizens (Ojambo, 2012). Between 1990 and 2019, Uganda saw a rapid increase in districts from 39 to 140 (Green, 2008; Mutebi et al., 2019). Despite its advantages in bringing services (e.g. outpatient health services, maternal and child health services, inpatient health services etc) close to people, district splitting has also negatively impacted service delivery capabilities, especially in health, by disrupting health system structures and creating new administrative burdens (Kuuml & Jane, 2014). Despite claims that splitting enhances service proximity, studies conducted elsewhere appear to suggest that it mainly serves political interests (Awortwi & Bert Helmsing, 2014). The consequences of such splits on health systems, particularly on workforce organization, remain underexplored.

In Uganda, health system decentralization paralleled political decentralization, devolving health system functions to local governments (Ojambo, 2012). This shift empowered district service commissions to manage human resources, including health workforce recruitment (Mangwi Ayiasi et al., 2019; Mansour et al., 2022; Uganda government, 1997). The tiered district health system, aligned with administrative structures, comprises various levels of health service provisions and mandates which corresponds to the workforce structures.

### The district health system structure and health workforce

The health workforce is intricately linked to the hierarchical decentralized political system, with each administrative unit having attached to the tiered healthcare levels and staffing norms (Green, 2008; Ramadhan, 2014) See table 1. District splitting directly affects the subnational health workforce structure and functionality, yet research primarily focuses on decentralization’s overall impacts (Bossert & Mitchell, 2011). This study aimed at using the chaos and systems theories to explore how the health workforce as a building block of the district health system re-organised itself following district splitting in Uganda.

### Theoretical Framework

The study utilized Chaos and Systems theories to examine the impact of district splitting on the subnational health workforce. Systems theory views organizations as interconnected subsystems, emphasizing holistic understanding (Chikere & Nwoka, 2015). Districts, comprising various subsystems including health, experience destabilization and subsequent system responses post-split.

Chaos theory suggests that system responses are nonlinear and unpredictable, characterized by self-organization and feedback loops (Levy, 1994; Rickles et al., 2007). This leads to disruptions and reorganization within health subsystems (Harrington & Voehl, 2016). Systems theory informed the conceptualization of health workforce organization, while chaos theory analyzed system response patterns post-split. The major components of chaos theory include; interconnectedness, feedback loops, and self-organization, evident in informal feedback processes and meetings facilitating efficient task performance and results attainment. These were applied to guide the analysis of the study.

### Study design

This was a cross-sectional exploratory study that used qualitative research methods with both deductive and inductive thematic analysis.

### Study sites

The study was conducted in four broader regions of Uganda i.e. Northern, Western, Eastern and Central. Within each of these regions, two districts were selected for inclusion into the study. Table two shows the eight districts selected for the study i.e. four old districts that existed before the split i.e. Lira, Pallisa, Mukono and Mbarara from which the new districts of Dokolo, Kibuku, Buikwe and Ibanda were curved out during the 2010 split. At least two districts i.e., an old and new were selected from each region hence representation from four regions of the country. The selection criteria of the four old districts were based on the Ministry of Finance and Economic Planning (MOFEP) and Development Social Economic Status (SES) (GoU, 2010; Heller et al., 2010; Kavuma Richard, 2009) where upon two districts were perceived to be performing well and the other two perceived to be performing poorly.

### Study participants

The study included a purposive sample of 22 participants: four elected district political leaders, five local government staff, four district service commission members, four district health team officials, and five district senior management members. Participants were selected based on their involvement in health workforce organization before and after district splitting, ensuring they were employed in the district during and after the split. Only those in leadership roles or governing committees responsible for health workforce organization were included, with equal representation from old and new districts. They were chosen for their strategic roles in health workforce planning and management.

## Data collection methods

### Key informant interviews

Interviews were conducted in December 2018, and the information elicited covered respondent’s experiences spanning fourteen years from 2005 to 2018. These were conducted with senior district officials involved in the recruitment process during district splitting. Participants were selected from various categories: District Service Commission (DSC) members (Administrative), DSC technical members (Administrative), District Health Managers (Technical), District Technical Leaders (Administrative local government), and District political leaders (Politicians). These categories were chosen for their critical roles in district planning and health workforce leadership. Participants from both old and new districts were included to enable comparison.

Experienced interviewers with master’s degrees conducted the interviews using a specially developed guide covering the major themes from the research questions. Each interview lasted about 45 minutes and was recorded digitally. Interviews took place at respondents’ preferred locations, usually their workplaces. The study also involved reviewing documents to understand local government policies and decentralization regarding DS. Key documents reviewed included the Public Service Standing Orders and the Human Resource for Health Policy in Uganda, which informed recruitment and deployment processes.

### Data analysis

Interview data were transcribed verbatim from audio recordings into Microsoft office word with transcripts were coded deductively following the applied framework domains in Atlas.ti version 7 where the codebook was developed and the codes reflected the chaos theories as an analytic framework, four themes were identified for deductive thematic analysis which included;: Systemic assignment of health workers based on location during DS, inter-district health worker migration (on secondment or appointment), intra-district health worker migration (within the district), and health workforce shortages (due to vertical and horizontal migration). These themes were analyzed to provide interpretive explanations, comparing data between old and new districts and different healthcare system levels at the subnational level.

The first author and a social scientist independently reviewed and coded interviews using a deductive thematic approach based on the key themes and later inductively following emerging insights from the datasets. They discussed individual matrices, focusing on coding differences until when variations were resolved. This process enhanced the quality, depth, and interpretation of results.

### Ethical considerations

The study was approved by the Institutional Review Board at Makerere University School of Public Health (HDREC 140) and the Uganda National Council of Science and Technology (SS 2678). Administrative clearances were obtained from district political and health sector leaders before conducting interviews. Respondents were informed about the study’s objectives, procedures, risks, and benefits. Interviewers had no prior contact with respondents. Interviews were conducted only after obtaining written informed consent and a separate consent for recording. They were conducted in quiet, secure locations chosen by respondents, interviews ensured privacy and confidentiality. All identifying information was anonymized and replaced with pseudonyms.

## Results

Overall, twenty-two respondents were selected from the eight districts with an average of three per district. The selection took into consideration the five types of cadre identified for interview.

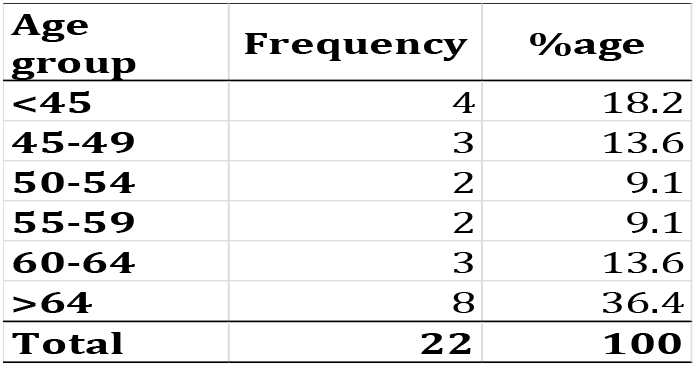

| Age group | Frequency | %age |
| --- | --- | --- |
| <45 | 4 | 18.2 |
| 45-49 | 3 | 13.6 |
| 50-54 | 2 | 9.1 |
| 55-59 | 2 | 9.1 |
| 60-64 | 3 | 13.6 |
| >64 | 8 | 36.4 |
| **Total** | **22** | **100** |

Out of the 22 respondents interviewed, those 64 years and above group (36.4%) had the highest number of respondents followed by those below 45 years (18.2%), 45-49 years (13.6%) and 60-64 years (13.6%) respectively. Age was generally balanced between old and new districts with almost equal numbers in each age group. There was only one female among the 22 respondents interviewed, which could be a sign that the most dominant top leadership positions are held by males. Using systems and chaos theory, the study examined the impacts of district splitting. Four themes emerged: random health worker assignment, inter-district migration, intra-district movement, and resulting shortages. Splitting the health system caused workforce reorganization, random worker allocation, and exacerbated shortages. This affected all administrative levels, disrupting operations and service delivery, and causing overall disruption, disorganization, and delayed stability in both old and new districts.

### Emergence of district health workforce management structures after reorganisation Systemic assignment of health workers

District splitting occurred along geographical boundaries defined by the local government act. Before splitting, health workers were attached to particular health facilities by the district health officer and chief administrative officer. Post-splitting, health workers were systemically assigned to new districts based on their location at the time of the split. Most workers remained at their stations, with few moving to support new districts. Those who moved left gaps, prompting self-organization to fill them. Staff mostly stayed put, with only seconded workers relocating, some later choosing to move permanently.

> *Most of our staff remained at their work stations after the districts split. It’s only those that were seconded to support the new district that moved, though some later made up their mind to move on to the new district (CAO 3)*.

### Inter-district migration of health workers

During district splitting, old districts seconded health workers to new ones as per MOLG/PSC requirements. Both old and new districts began appointing staff to fill vacancies. Seconded staff, officially sent by old districts to new ones, started work while recruitment progressed. This led to gaps in old districts, necessitating redeployment. Meanwhile, seconded staff were free to apply for relocation or higher positions. Old districts retained key personnel, often senior staff, influencing new district staffing. Tensions arose when new districts rejected non-native staff, requiring higher authority intervention. Informal negotiations and lobbying formed to resolve disputes, promoting self-organization. Over time, tensions eased through reconciliation, showcasing the interconnectedness and power of dialogue among involved parties.

> *We in the old district had the advantage of choosing who goes to the new district especially when it came to setting it off, but still HWs who wanted to move due to varying reasons, still moved on whether we accepted or not and we could not stop them from moving. So those were struggles under the carpet that were agreed on just like that (LC V Chair 1)*.
>
> *Seconding health workers was not an easy task because either district wanted the best. I remember how the new district rejected the person we sent them and demanded for another person. To me a beggar has no choice. You either take what is available or leave (CAO 3)*.

During the secondment process, friction arose as districts competed to attract senior or highly qualified health workers. Some districts resisted releasing staff, while new districts offered attractive terms of engagement/deployment, leading to unauthorized staff movements and chaos. Individual decisions escalated, involving district leadership and triggering the “butterfly effect”. In some instances, reconciliation efforts, with support from Health Service Commission (HSC) members and local opinion leaders, restored order through reorganization and return to sanity within the districts.

> *“You know time came when we competed for the good health workers we knew. We had to offer better terms than our competitors so that the health worker is convinced to join our district. This is allowed in this new arrangement of decentralisation” but in most cases there was a lot of unofficial agreements between the administration and staff (CAO 2)*
>
> *We witnessed a lot of behind the screens moves as some health workers were lobbying for movement to another district, and others were negotiating for promotion even before being appointed. But because we are not technical, we let them do the negotiation and the technical personnel handled that (LC V Chair 2)*.

The secondment and appointment process received utmost attention across districts, with District Service Commissions (DSCs) resisting any external influence. However, districts prioritized qualified natives over “external” professionals with similar qualifications, causing tension among leaders and necessitating crisis meetings. Feedback and consultations between political and technical leadership arms, along with involvement of Health Service Commission (HSC) or Public Service Commission (PSC), led to consensus on the way forward, restoring normalcy.

> *Let me tell you, the decision to retain and refer HWs created a lot of tension with a lot of lobbying by both district councillors, HWs and the technocrats. At one point we were in a planning meeting and some members started accusing each other of influencing the screening to the extent of almost exchanging blows (DSC Chair 3)*.
>
> *You know these district councillors thought they could force me into appointing people without the right qualifications, I refused and assured them I was not willing to face the inspector general of government because of corrupt tendencies in the recruitment. I even asked them to put it in writing and I forward it to our seniors in the ministry. That’s when they all went silent (CAO 5)*.

In new districts, District Health Officers (DHOs) served in acting capacities after being seconded from old districts or appointed by the public service commission to initiate district health offices.

Many seconded health workers assumed automatic recruitment leading to preferential treatment of health workers from old district which could have led to not following recommended procedures when recruiting staff. This disorganized relations between workers and leaders, impacting inter-district cooperation. Negotiations, mostly informal, resolved disagreements, but in cases of irreconcilable differences, health workers returned to previous districts or sought employment elsewhere.

> *“When I was seconded to the new district, I thought it was obvious they were going to recruit me because I had all the qualifications but surprisingly, I started hearing talk from some colleagues that I was not a son of the soil. Meaning that because I was not born in the district, the DSC had some other person they preferred. I was only saved by the HSC representative who insisted that the person they had fronted did not qualify for the post*.*” (DHO 2)*

The final task of appointing and deploying after secondment faced disruptions and interference from various leaders. District Service Commissions (DSCs) and District Health Officers (DHOs), guided by acting Chief Administrative Officers (CAOs), were responsible. Interference affected DHOs, as health workers requested deployment near their homes or towns. DHOs would occasionally hold informal meetings before releasing deployment schedules, involving district leaders and technocrats to maintain good relations. Acting positions in new districts made technical leaders vulnerable to political pressure, affecting decision-making. Appointments often occurred in the presence of a Health Service Commission (HSC) member, providing support to technical personnel.

*In my district we had several qualified health workers who had been in the same positions for several years without being promoted due to lack of opportunities. We therefore considered them during the recruitment process and appointed them on probation in new positions as a way of motivation. (CAO 3)*.

Many seconded staff from old districts were appointed on probation in their new districts, while some acting staff not meeting requirements were advised to return to their previous districts or accept lower positions, causing distress and disorganization. Those seeking official transfers but not accepted faced dilemmas, as returning to previous districts was often not feasible, forcing them to seek employment elsewhere. There was different treatment/handling of health workers on a case by case basis which is reflective of lack or abuse of standard operating procedure for managing human resource for health.

> *“I didn’t like the way some of the seconded staff were treated by the DSC just because they preferred candidates that were born in this district. We lost a chance of getting very experienced and qualified staff because the DSC wanted people born in the district “son of the soil”. (DSC Sec 2)*

### Intra district migration of health workers

Following district splitting, health worker movement occurred within both old and new districts due to gaps left by those on secondment or seeking new appointments elsewhere. Both districts identified gaps, advertised vacancies, conducted interviews, and appointed new staff to fill the gaps. Meanwhile, reorganization of available workforce ensured continued service delivery despite gaps. District Health Officers (DHOs) redeployed staff to facilities with vacancies, facing challenges as some workers resisted relocation. Meetings between health workers and their leadership were held to resolve disputes, sometimes involving Chief Administrative Officers (CAOs). Redeployment occurred in all districts post-splitting to address workforce gaps. After probationary appointments, district health offices reorganized staff at all levels to maintain quality healthcare services.

> *Immediately after DS occurred, we had to do reallocation of health workers to fill some of the key positions that had been created due to seconded staff. Then we also reorganised the workforce after the final appointment of new staff (CAO 1)*.

### Health workforce shortages

Throughout the various forms of reorganization, the existing health workforce was distributed across districts, creating gaps in each district, doubling the pre-splitting workload. Recruitment efforts failed to fill all gaps, leading to further disruptions. Emergent gaps due to seconded or unauthorized departures required reorganization and advertising of vacancies, exacerbating understaffing and disrupting health facility operations. Districts already grappling with understaffing pre-splitting faced worsened conditions. Filling vacancies awaited confirmation from the Ministry of Finance, Planning, and Economic Development (MOFPED) on the wage bill. Constant feedback meetings among affected district leaders facilitated secondments and redeployments, achieving workforce balance across districts before addressing persisting gaps through advertisement and recruitment.

## Discussion

This study aimed at using the chaos and systems theories to explore how the health workforce as a building block of the district health system re-organised itself following district splitting in Uganda. This qualitative study examines the reorganization of the health workforce following district splitting in Uganda, revealing its impact on both new and old districts. It explores how reorganization within and between districts results in multidirectional workforce arrangements, such as systematic assignment of health workers, inter-district migration of health workers, intra district migration of health workers and health workforce shortages. Subsequently, these will be elaborated on further in the sections that follow. The study offers fresh insights into the interconnectedness of the health workforce and district systems, shedding light on the complexities of their reorganization after district splitting.

The study also critiques broad discussions on decentralization, urging for a deeper understanding of local government systems and their subsystems(Zaidi et al., 2019) to inform health worker allocation following district splitting. It stresses the importance of policymakers comprehension of these subsystems’ interactions and formations to address the positive and negative impacts of district splitting effectively(Abimbola et al., 2019). Despite challenges like respondent recall, the study utilizes triangulation of data sources to mitigate these issues, enhancing the reliability of its findings(Beesley et al., 2011; Durham et al., 2015). Detailed discussions about the results following themes from the applied framework follows here next;

### Systematic assignment of health workforce

Before district splitting, the decentralisation policy through the local government act gave districts autonomous powers to advertise, interview and recruit health workers. These health workers were already stationed across health facilities in all the selected districts. However, after the splitting, most health workers remained at their original workstations based the district appointment and deployment plans under their local government demarcations. Many avoided moving due to observed disruptions and chaotic scenarios. Some were content with their current stations, while others were reluctant to undergo the rigorous process of switching districts, which involved either secondment by the Chief Administrative Officer (CAO) or applying through the standard District Service Commission procedures. Another study conducted in Kampala city showed that those who have lived in rural areas were less likely to report wanting to emigrate(Nguyen et al., 2008).

### Inter-district migration of health workers

After district splitting, some health workers were seconded to new districts by Chief Administrative Officers (CAOs) to kickstart operations, causing disruptions in the old districts. This led to gaps due to staff departures on secondment or appointment. New positions were created at different levels in the health system structures of both new and old districts. However, the movement of health workers between districts was not smooth, disrupting both subsystems and the entire district structure. Disagreements arose on how to share available human resources, affecting recruitment components interconnected within the system. In other studies it was noted that these disruptions triggered emergent behaviours and took time to reorganize, mainly due to political and technical differences between districts(Chen et al., 2021; Nalukwago et al., 2019).

In this study, negotiations and rule bending occurred during secondments and appointments, breaking some rules in order to facilitate recruitment. In a similar study the tension between rule makers and breakers influenced the ecology of the field(Dzansi et al., 2016). Similar findings from another study showed that negotiations, bargaining, and conflicts between district leaders, technocrats, and health workers ensued, leading to disorganization within and between districts(Parrish, 2022). However, through feedback negotiations, the chaos was eventually normalized into a reorganized state(Renn, 2022; Schein, 2004).

In this study, a lot of negotiations during this period of splitting brought out the interconnectedness of different players and self-organization which made it possible for agreements to be reached. This chaotic period highlights the dynamics between rule breakers and makers, shaping both tight and loose cultures within organizations(Lithwick, 2012). While chaos presents challenges, it also leads to innovative solutions(Gelfand, 2019; Parrish, 2022). Studies in Kenya have shown recruitment strategies focusing on geographic areas to minimize staff transfers(Adano, 2008).

### Intra-district migration of health workers

The recruitment process faced internal and external disruptions, delaying the process in most districts. Suspected ulterior motives among committee members led to delays, requiring extensive discussions and informal meetings for resolution. Respondents considered these to be a result of political interference by different players which later resulted into several informal meetings and consensus was reached by all concerned parties. Therefore, the formal process was compromised, prompting management to adopt informal transparency measures, settling disputes and allowing work to proceed. In another study it was seen that the decision-making in behaviour change is often quantum rather than linear, with changes being conceptualized as deterministic processes(Kenneth Resnicow, 2008). It was also observed that small issues during redeployment caused friction among leaders but were resolved through formal and informal discussions, reaching consensus. Some behaviour change events may be linear, while others may be complex and nonlinear, requiring a chaos lens for understanding(Ken Resnicow & Vaughan, 2011; Kenneth Resnicow, 2008).

### Health workforce shortages

After the completion of secondment, appointment, and redeployment across all districts, new gaps emerged due to unauthorized departures, requiring reorganization and advertising of vacant posts. These unforeseen movements disrupted work at health facilities, worsening understaffing and posing challenges in transferring recently deployed workers. Time constraints and financial implications compounded the urgency to fill vacancies before the end of the financial year. The complex and chaotic nature of health worker behaviour change can be integrated into public health interventions, offering insights into how individuals and organizations adapt(Gardner et al., 2021). Despite completing the recruitment process, new gaps arose from unauthorized departures, further disrupting the system. A complementary approach is suggested, encouraging the incorporation of nonlinear concepts into intervention design and analysis(Bien, 2004; C’De Baca & Wilbourne, 2004; Miller, William R C’de Baca, 2001; Miller, 2004). The solutions of yesterday had now become the problems of today (Agyepong et al., 2012). Support from Chief Administrative Officers (CAOs) is crucial for District Health Officers (DHOs) during deployment to mitigate complaints and discontent among health workers.

The study underscores the implications for reconceptualizing workforce management in decentralized settings post-district splitting to enhance service delivery efficiency at sub-national levels, especially in human resource for health organization. It challenges the instrumentalist view of district creation as merely bringing services closer to the population, revealing unintended chaos in health workforce distribution. By employing a chaos lens, the study highlights non-linear disruptions and disorganization within the health workforce subsystems, emphasizing the need for a more systematic approach to decentralization and district splitting to achieve efficiency goals(Zanin & Martínez, 2022).

### Strengths and limitations of the study

This paper is informed by district leaders and health workers perspectives of what happened with the reorganisation processes after districts were split. Their views inform our conclusions about the effect of district splitting human resources for health management focusing on the reorganisation processes. All findings are from people who were present and directly participated in these processes, the study brings first-hand experiences of what transpired, while the systems and chaos theories show us the unforeseen inefficiencies that occurred in the reorganisation process after splits had occurred. Its conclusions can reliably be assumed in respect of other districts that have experienced district splitting. There could have been some social-desirable bias given the area under study. It’s possible that there was some recall bias given that the study was conducted some years after district splitting of 2010. It’s also possible that there was social desirability and not wanting to talk about bad things about the district and its human resource by some respondents. Due to the cross-sectional nature of the research, it was unable to observe and provide for analysis of trends over time. Findings from the few study districts are hard to generalise as experiences may have varied in other districts hence impeding generalisability.

## Conclusions

District splitting affected all district systems right through the physical divide along geographical boundaries, through the whole district administrative structure, splitting the district councils, district local government (all departments including health), DSCs etc. The health system was generally affected as it aligns with the district local government structure from top to bottom, and is categorised into four sub systems. These subsystems are interconnected, interrelated, and interdependent, working together to produce optimal outcomes while collectively striving to reorganize the health workforce to deliver quality healthcare.

We see how the formal system with linear thinking is disrupted and disorganised by political and technical interference before it can reorganise itself back to normal. Even when the districts think they are done with the reorganisation, and gaps have been filled, then they realise that some health workers left their stations without official release. Then the process of reorganisation continues on, meaning that the solutions of yesterday become the problems of today.

Findings suggest that without considering the consequences of underlying patterns, interconnected actions, continuous feedback loops, and self-organizing mechanisms after splitting, systems and subsystems do not perform as expected. Unexpected small errors can arise, leading to a significant negative impact on the health workforce reorganization process.

The chaotic nature involves disorder, disruption and disorganisation of systems including the health workforce. But then the process of reorganisation that follows chaotic happens, introduces enabling conditions that include innovation, creativity and flexibility. These introduce an environment of understanding among the different district leaders and health workers. If these are considered during the planning and implementation of district splitting processes, they are likely to benefit both parties and reduce the tension or chaos that is caused by the splitting.

## Data Availability

No legal or ethical concerns

## Declarations

### Ethical approval and consent to participate

This study has received ethical approval from Makerere University School of Public Health Higher Degrees Research and Ethics Committee (HDREC) ref; HDREC 140. The final ethical clearance is being processed from the Ugandan National Council for Sciences and Technology (UNCST) ref; SS 2678. Further administrative clearance was obtained from the management of all participating health facilities and organisations where the study was conducted. Individual informed consent was obtained from all prospective respondents.

### Consent for publication

Consent to publish the findings of the study was obtained at the point of seeking consent to participate. The participants’ confidentiality and anonymity while reporting was assured.

## Not applicable

### Availability of data and materials

The datasets used and/or analysed during the current study are available from the corresponding author upon reasonable request.

## Author’s contribution

AM conceived the study; ER and FM provided supervision support. MM guided the methods with special attention to the theories and SK helped with conceptualisation of health systems. All authors reviewed and approved the final manuscript.

## Acknowledgements

The author wishes to acknowledge the contribution of the data collection team and the reviewers who provided useful insights that informed the revision of the manuscript.

